# Traumatic Experience and Self-Control in Old Age

**DOI:** 10.1101/2022.03.21.22272686

**Authors:** Youngjoo Choung, Tae-Young Pak

## Abstract

The behavioral economics literature suggests that exposure to traumatic events shifts preference features like risk aversion and time preference. Drawing on this literature, this study explored the relationship between early life exposure to traumatic events and self-control at older ages. The data were obtained from the Health and Retirement Study, which offers retrospective data on trauma exposure and a measure of self-control. The results showed that the experience of serious physical attacks or assaults was associated with a 3.1% reduction in self-control, above and beyond the influence of demographic characteristics and childhood socioeconomic disadvantages. The mean number of years elapsed since the physical attack was about 30, conditional on exposure, indicating that traumatic experiences early in life exert a lasting influence on self-control. Our findings were consistent with evidence that experiences of natural disasters and armed conflicts increased impatience among survivors.

**JEL classification code:** D12, D14, D91

## 1. Introduction

The predictive validity of self-control for adult life outcomes has been demonstrated in a variety of domains. For instance, pre-school children who could wait longer for their preferred rewards in a controlled experimental setting developed into more academically and socially competent adolescents, achieving higher SAT scores and coping maturely with frustration and stress (Mischel et al., 1988; Shoda et al., 1990). In follow-up research, this self-control test proved to have significant predictive validity for educational and health outcomes over the life span (Moffitt et al., 2011). Economists have modeled self-control as a parameter of “willpower” in the intertemporal choice model (Thaler and Shefrin, 1981) and have demonstrated its association with savings behavior, occupational achievements, and financial well-being (Daly et al., 2015; Kaur et al., 2015; Laibson, 1998; O’Donoghue and Rabin, 2000; Strömbäck et al., 2017).

Numerous studies have investigated why some individuals are better at self-control than others. This research stream has revealed significant individual heterogeneity across various demographic and socioeconomic characteristics (Calero et al., 2007; Cobb-Clark et al., 2019; Silverman, 2003; Matthews et al., 2009). Much of this heterogeneity has been attributed to differences in parental and family environments that could determine the socialization process during childhood (Gottfredson and Hirschi, 1990). However, this claim has been challenged by the finding that measures of parenting have little explanatory power for self-control in empirical analyses (Beaver, 2009). Some studies have found that parenting is no longer a significant determinant of self-control once the model accounts for genetic factors and prenatal conditions (Beaver et al., 2009; Wright and Beaver, 2005).

It is likely that other salient factors that could determine the learning of self-control have not been identified by the literature. This study focuses on the lasting negative influence of traumatic experiences on self-control. The human brain is known to encode external stimuli as threatening or non-threatening and to adjust its behavior to the perceived threat level (McEwen, 2007). When experiencing a potentially threatening event, it enters a state of “mental combat mode” and redirects energy and resources away from advanced reasoning and toward a lower level of basic bodily processes necessary for survival (Arnsten et al., 2015). Sustained exposure to threats and the related hormonal changes have lasting effects on brain architecture; these lead to an under-activation of the brain’s modulatory functioning and thus undermine its ability to resist temptation (National Scientific Council on the Developing Child, 2005).

Most studies on traumatic experience and impulsive behavior have examined adolescents or young adults, focusing on the role of domestic violence and childhood abuse in the socialization process. As a result, current knowledge is limited to the relatively short-term effects of traumatic events experienced in early childhood (Simmen-Janevska et al., 2015). The question of whether shifts in self-control persist into later life has significant implications for economic behaviors and poverty dynamics, given their salience for predicting later life outcomes. This study examines the association between traumatic life experiences and self-control in older adults using retrospective data drawn from the Health and Retirement Study (HRS). Our results show that the experience of serious physical attacks or assaults is associated with about a 3.1% decline in self-control up to three decades later. The estimated trauma effect is roughly comparable to the effect of being unemployed and three times greater than the gender gradient in self-control, indicating that early life trauma is a major determinant of self-control with lasting consequences.

This study contributes to three strands of research. The first is research on early life experiences and human capital accumulation, which reports the association between adverse conditions (e.g., illness, in-utero alcohol exposure, poor nutrition) during early childhood (the period from in-utero to age five) and health and economic outcomes in later life (Almond and Currie, 2011; Ampaabeng and Tan, 2013; Blackwell et al., 2001; Hayward and Gorman, 2004). Although self-control is generally not considered a form of human capital, it is a valuable psychological resource that enables people to follow through on their commitments to human capital investments and expedites their accumulation (Cadena and Keys, 2015; Griesdorn and Durband, 2016).

Our work also adds to the growing body of literature that estimates the effect of traumatic experiences on preference features, such as risk aversion and time preference. Evidence that preferences shift after exposure to a traumatic event has been reported for survivors of major natural disasters (Ahsan, 2014; Callen, 2015; Cameron and Shah, 2015; Cassar et al., 2017; Eckel et al., 2009; Hanaoka et al., 2018), terrorist attacks (Sacco et al., 2003), and civil conflicts and wars (Bellucci et al., 2020; Brown et al., 2019; Callen et al., 2014; Jakiela and Ozier, 2019; Kim and Lee, 2014; Moya, 2018), exploiting the event as a source of identification for the trauma’s causal effect. A similar finding has also been reported in a study of retrospective data on negative life events and risk preference (Banks et al., 2020; Bucciol and Zarri, 2015; Hetschko and Preuss, 2020). Despite this mounting evidence, however, the direction of the preference shifts remains unclear, with the findings differing according to the study site, source of traumatic events, and preference outcomes examined (Abatayo and Lynham, 2020).

Finally, our study addresses the literature in psychology and human development that explores how self-control is formed and evolves over a lifetime. The conventional theory has been that self-control is established early in life and remains relatively stable afterward (Gottfredson and Hirschi, 1990). Hence, current knowledge is unable to help us determine why some individuals continue to exhibit impulsive behaviors long after exposure to toxic stress (e.g., among veterans home from war) and what this implies for their long-term economic well-being. We depart from this conventional practice by taking a longer, lifetime perspective of self-control development.

The remainder of the paper is organized as follows. Section 2 reviews the research that links traumatic experiences to self-control and explains how this issue has been modeled in studies of economic preferences (e.g., risk aversion, time preference). Section 3 describes our data, measures, and empirical models. Section 4 presents our estimation results. Section 5 concludes with a discussion of the study’s findings and implications, and potential future research directions.

## 2. Previous research

### 2.1 Traumatic experience as a determinant of self-control

The Diagnostic and Statistical Manual of Mental Disorders defines trauma as exposure to “actual or threatened death, serious injury, or sexual violence” (American Psychiatric Association, 2013). Common experiences of trauma include direct exposure to trauma, witnessing trauma, or learning about a traumatic event that happened to a family member or close friend, in the form of abuse or neglect, accidents, armed conflict, domestic violence, and natural disasters (De Bellis and Zisk, 2014). A memory of childhood trauma is known to increase the risk of post-traumatic stress disorder and leads to depression, antisocial behaviors, and substance abuse (Hovens et al., 2012; Yehuda et al., 2001).

A growing body of psychiatry research is examining the link between trauma exposure and self-control problems. Studies have found a positive association between trauma in childhood and pathological impulsivity in patients with borderline personality disorder (Figueroa and Silk, 1997), gambling disorder (Ledgerwood and Petry, 2006), substance abuse (Weiss et al., 2012), and suicidal behavior (Roy, 2005). Even in healthy populations, the memory of abuse or neglect encountered early in life has been found to be related to reduced executive functions in adulthood (Spann et al., 2012), deviations in cognitive control (Rogosch et al., 1995), and poor self-regulation over smoking and drinking (Goldstein et al., 2013). Psychological responses to adverse experiences and the associated emotional effects have also been found to be potential paths leading to executive dysfunction (Duckworth et al., 2013).

Experiencing trauma can have an adverse impact on the brain’s modulatory functions. When the body senses environmental threats, the stress response system becomes activated and shifts the body’s metabolic resources away from the thinking part of the brain and toward more primitive functioning related to survival (De Bellis and Zisk, 2014). While the body is fighting off potential threats, advanced brain functions not necessary for survival become less active. As a result, the frontal lobe—the decision-making center of the brain—starts functioning slowly, and its connectivity with other parts of the brain is weakened. This process helps individuals mobilize more effectively in the event of an emergency but dampens their ability to make complex decisions such as controlling impulses (Marshall et al., 2016).

Some studies have found that trauma leads to a permanent change in brain structure (De Bellis and Zisk, 2014). Neuroimaging studies have shown that childhood trauma is associated with reduced volume in the brain areas that manage memory/emotion processing and behavioral regulation (Hoy et al., 2012). Similarly, pediatric research has demonstrated that the volume of certain brain areas related to executive functions is smaller in abused and deprived youths than in non-maltreated youths (Bauer et al., 2009; De Bellis and Kuchibhatla, 2006). Experiments with animals have also shown that external stress impairs the performance of tasks requiring top– down decision making and triggers structural changes in brain circuits (Arnsten, 2009). Whether such changes in brain structure persist into old age is currently unknown.

### 2.2 Traumatic experience and economic preferences

Our study is broadly related to the research arguing that negative life experiences may alter economic preferences, such as risk aversion and time preferences. These studies have investigated unexpected onsets of historical events (e.g., natural disasters, armed conflicts, economic crises) as a source of trauma and have examined the extent to which such events correlate with survivors’ preference features. For instance, Cameron and Shah (2015) conducted risk game experiments with Indonesians living in disaster-prone areas and found that those who had experienced floods or earthquakes tended to choose less risky options. Similarly, Jakiela and Ozier (2019) examined the post-election crisis in Kenya and related civil conflict and found higher risk aversion of about 80% among those who were directly exposed to the conflict. An upward shift in risk aversion after experiencing a traumatic event has also been confirmed in survivors of cyclones (Ahsan, 2014), tsunamis (Cassar et al., 2017), drug-related violence (Brown et al., 2019), and wars (Belluci et al., 2020; Callen et al., 2014; Kim and Lee, 2014; Moya, 2018). In particular, Cassar et al. (2017) examined a measure of time preference along with their analyses of risk aversion and found that the 2004 tsunami in Thailand caused its victims to be more impatient.

The fact that these studies were conducted a few years or decades after the events occurred seems to indicate that the preference shift is long-lasting, though perhaps not permanent. Especially noteworthy are Belluci et al. (2020) and Kim and Lee (2014), who showed that exposure to war is linked to risk preference several decades later. The former study used childhood exposure to war as a natural experiment and found that those who lived in a region with intense battles during the war exhibited higher risk aversion at older ages. Kim and Lee (2014) found that the association between war intensity and risk aversion was more pronounced for those aged four to eight during the war, suggesting that experience during the period most sensitive for risk attitude formation has a more lasting influence. Bucciol and Zarri (2015) yielded a similar finding through analyses of retrospective data on past life experiences, in which the experience of a physical attack or losing a child was found to crowd out risky investments from the family financial portfolio.

The question regarding the direction of the preference shift remains unsettled in this literature, with some of the evidence suggesting that preferences altered in the opposite direction after the event. Studies like Page et al. (2014) and Kahsay and Osberghaus (2018) found a downward shift in risk aversion among survivors of flood and storms, while Abatoyo and Lynham (2020), Eckel et al. (2009), and Hanaoka et al. (2018) showed that the risk-seeking response after natural disaster is significant only for a particular gender. Voors et al. (2012) found increased impulsivity and altruistic behavior in the aftermath of a civil war, but Callen (2015) found an increase in the monthly discount factor among tsunami survivors in Sri Lanka.

## 3. Methods

### 3.1 Data description

Our data are drawn from the HRS, a large-scale longitudinal survey conducted every even-numbered year since 1992 by the Institute for Social Research of the University of Michigan. The HRS covers Americans over age 50 living in the United States. It is a rich source of data on the changing life circumstances Americans undergo toward the end of their work lives and in the years after retirement. The HRS covers four broad topic areas: income and wealth, labor supply, health, and healthcare access. The measures that are key for our study, self-control and traumatic experiences, were added to the HRS in 2006 as a part of the Psychosocial Leave-Behind (LB) module. The LB module was administered to 50% of the HRS respondents on a rotating schedule, so that each one responded to the module every four years. Traumatic experiences were fielded in the 2006, 2008, 2010, and 2012 HRS, while self-control was fielded in the 2010 and 2012 ones.

Our study sample consists of respondents aged 50 to 80 in the 2010 and 2012 HRS. It is restricted to those who answered questions on self-control and traumatic experiences, as well as other miscellaneous measures that are included in the regression as control variables. The final sample comprises 8,767 person-level observations (4,714 from the 2010 survey and 4,053 from the 2012 one).

### 3.2 Measure of self-control

Self-control was measured using a six-item scale taken from the Multidimensional Personality Questionnaire (Tellegen, 1982). Respondents were asked to rate their impulsive behavior in the domains of money management (Q1: “I keep close track of where my money goes”), attention to tasks (Q2: “I often stop one thing before completing it and start another”), action with deliberation (Q3: “I often act without thinking”), tendency to forecast (Q4: “Before I get into a new situation, I like to find out what to expect from it”), alertness (Q5: “I am often not as cautious as I should be”), and planning ahead (Q6: “I often prefer to play things by ear rather than to plan ahead”) on a six-point Likert scale, ranging from “strongly disagree” (1) to “strongly agree” (6). Negatively phrased items (Q2, Q3, Q5, and Q6) were reverse-coded so that a higher value of the summary score represents better self-control. A summary score is the mean of the six responses.

### 3.3 Measures of traumatic experiences

Traumatic experiences were assessed using the lifetime trauma inventory developed by Krause et al. (2004). The traumatic events considered in this study include (a) a serious physical attack or assault (“physical attack” hereafter); (b) a major fire, flood, earthquake, or other natural disaster (“natural disaster” hereafter); and (c) a life-threatening illness or accident (“life-threatening illness” hereafter). For each event, respondents were asked to indicate whether it had occurred to them and when it had happened last. Using these responses, we construct binary indicators of occurrence and of whether the event occurred before or after the threshold age of brain maturity. Approximately 7% of our sample reported a physical attack, 17% reported exposure to a natural disaster, and 25% reported suffering from a life-threatening illness. The mean number of years elapsed since the event is 30.3, 27.6, and 19.2 for physical attacks, natural disasters, and life-threatening illnesses, respectively (see Figure 1). The survey questions are provided in Table A1.

**Figure 1.**
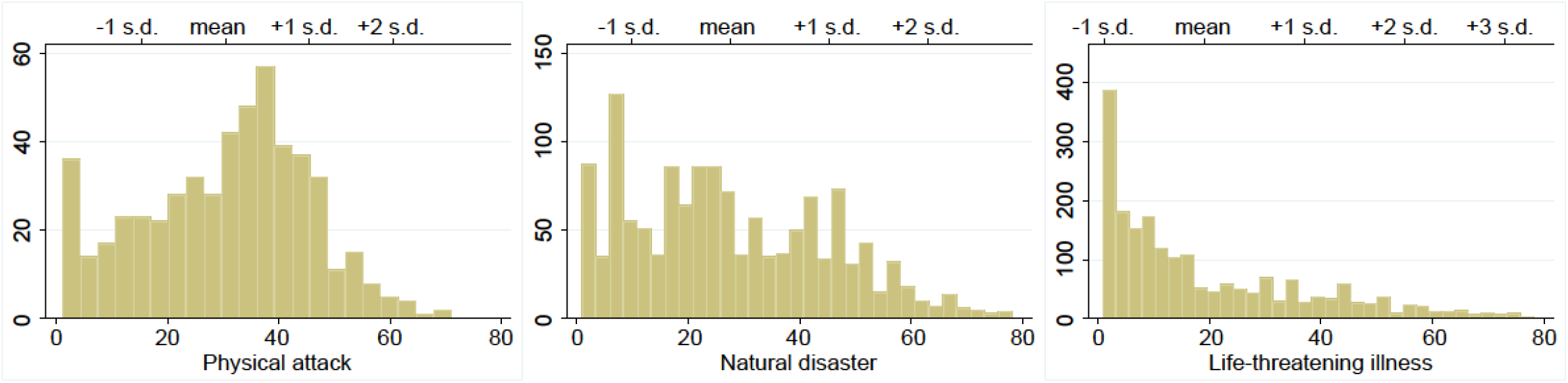
Frequency histogram of years elapsed after traumatic events

### 3.4 Empirical specification

We specify the following form of the regression for individual *i* at time *t ∈* {2010, 2012}:

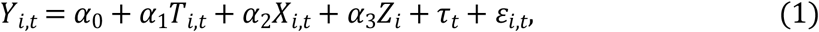

where *Y*_*i,t*_ is a six-point score for self-control, *T*_*i,t*_ are variables for the occurrence of traumatic events or the timing of their occurrence, *X*_*i,t*_ is a vector of covariates at time *t, Z*_*i*_ are indicators of early life conditions, *τ*_*t*_ are time fixed effects, and *ε*_*i,t*_ is an *i*.*i*.*d*. disturbance term. *τ*_*t*_ are dummies for the survey month and for the 2012 survey, which represents the average time effects that are common for individuals surveyed at the same time.

*X*_*i,t*_ includes age (in years), gender (female or male), race (Black or Hispanic, White, or other races), marital status (married or not in a marital relationship), number of living children, self-rated health (poor or fair, good or better), number of private health insurance plans, out-of-pocket (OOP) medical spending, employment status, household income, total net worth, and census region of residence (Northeast, Midwest, South, West, or other). The OOP medical spending is the amount spent on medical services by the respondent since the last interview. Employment status is coded with three exclusive categories: fully or partially employed, unemployed, and retired or not in the labor force. Household income is the sum of earnings, pensions and annuities, social security benefits, government transfers, capital income, and other income. Total net worth is the net value of financial and non-financial household assets, including residence, real estate, vehicles, business equity, and retirement accounts. Household income is transformed by *log*(*x* + 1) and total net worth by the log-modulus transformation. All dollar figures are in 2012 rates.

The respondent’s socioeconomic characteristics around the time he/she was exposed to the event is a potentially omitted factor that could confound the estimation of *α*_1_. Leaving early life conditions out of the model could lead to a biased estimation of *α*_1_ since they may partially determine the propensity to experience traumatic events and self-control in later life. To address this concern, we expand our regression with a vector of variables for the respondent’s characteristics in childhood and in mid-life. Specifically, *Z*_*i*_ includes variables for place of birth (born in the south or in other regions), parental educational background (years of education), quality of the relationship with parents before age 18, and history of parental substance abuse. It also includes OOP medical spending, employment status, household income, and total net worth, obtained from the 1998 survey (HRS, AHEAD, CODA, and WB cohorts) and the 2004 survey (EBB cohort). Table A1 presents detailed descriptions of these variables.

Regression models are estimated by a linear OLS. To provide a more intuitive interpretation of effect sizes, we present percentage change interpretations based on back-of-the-envelope calculations.

## 4. Results

Table 1 shows the average descriptive statistics for the study sample. The sample characteristics are presented for the full sample and for subsamples divided according to trauma exposure (a sample of 3,471 persons with an experience of physical attack, natural disaster, or life-threatening illness and a sample of 5,296 without). We find that the average respondent is 63.2 years old, has an average of 2.66 living children, owns 0.75 private health insurance plans, and holds a net worth of $559,858. Most participants are married (72%) and rate their health condition as good or better (81% good or better; 19% poor or fair). In terms of lifetime trauma, 7% report having experienced a physical attack, 17% report having experienced a natural disaster, and 25% report a life-threatening illness. Comparing the self-control scores by trauma exposure shows that those who experienced a traumatic event have lower self-control than those who have not (*p* < 0.05).

**Table 1.** Average descriptive statistics (*N*=8,767)

|  | No trauma<br>(5,296 obs.) | Trauma<br>(3,471 obs.) | Full sample |
| --- | --- | --- | --- |
| Self-control | 4.40 | 4.30<sup>a</sup> | 4.36 |
| Physical attack |  |  | 0.07 |
| Natural disaster |  |  | 0.17 |
| Life-threatening illness |  |  | 0.25 |
| Age | 63.00 | 63.58 | 63.22 |
| Female | 0.57 | 0.51<sup>a</sup> | 0.55 |
| Black or Hispanic | 0.11 | 0.10<sup>a</sup> | 0.10 |
| High school graduate | 0.33 | 0.29<sup>a</sup> | 0.32 |
| Some college | 0.24 | 0.31<sup>a</sup> | 0.27 |
| College and above | 0.34 | 0.34 | 0.34 |
| Married | 0.74 | 0.70<sup>a</sup> | 0.72 |
| No. of living children | 2.68 | 2.62 | 2.66 |
| Poor or fair health | 0.14 | 0.26<sup>a</sup> | 0.19 |
| No. of health insurance | 0.77 | 0.72<sup>a</sup> | 0.75 |
| OOP medical spending (\$) | 2735 | 3764<sup>a</sup> | 3137 |
| Employed | 0.45 | 0.37<sup>a</sup> | 0.42 |
| Unemployed | 0.04 | 0.04 | 0.04 |
| Household income (\$) | 101,409 | 89,523<sup>a</sup> | 96,760 |
| Total net worth (\$) | 585,069 | 520,614<sup>a</sup> | 559,858 |

*Notes:* Health and Retirement Study, 2010-2012. Sample characteristics are weighted by individual- and household-level weights provided in the RAND HRS data. Dollar figures are adjusted to 2012 dollars using the Consumer Price Index for all urban consumers (CPI-U). <sup>a</sup> Two-sample *t*-test by traumatic experience rejects the null hypothesis of no difference at the 5% significance level.

Table 2 presents the main regression results. We begin with the null model controlling for only trauma indicators and gradually expand the model with additional control variables. Coefficient estimates for the month-of-survey dummies, year-of-survey dummies, and census region fixed effects are omitted for brevity. All standard errors are clustered at a combination of individual and age levels (two-way clustering).

**Table 2.**
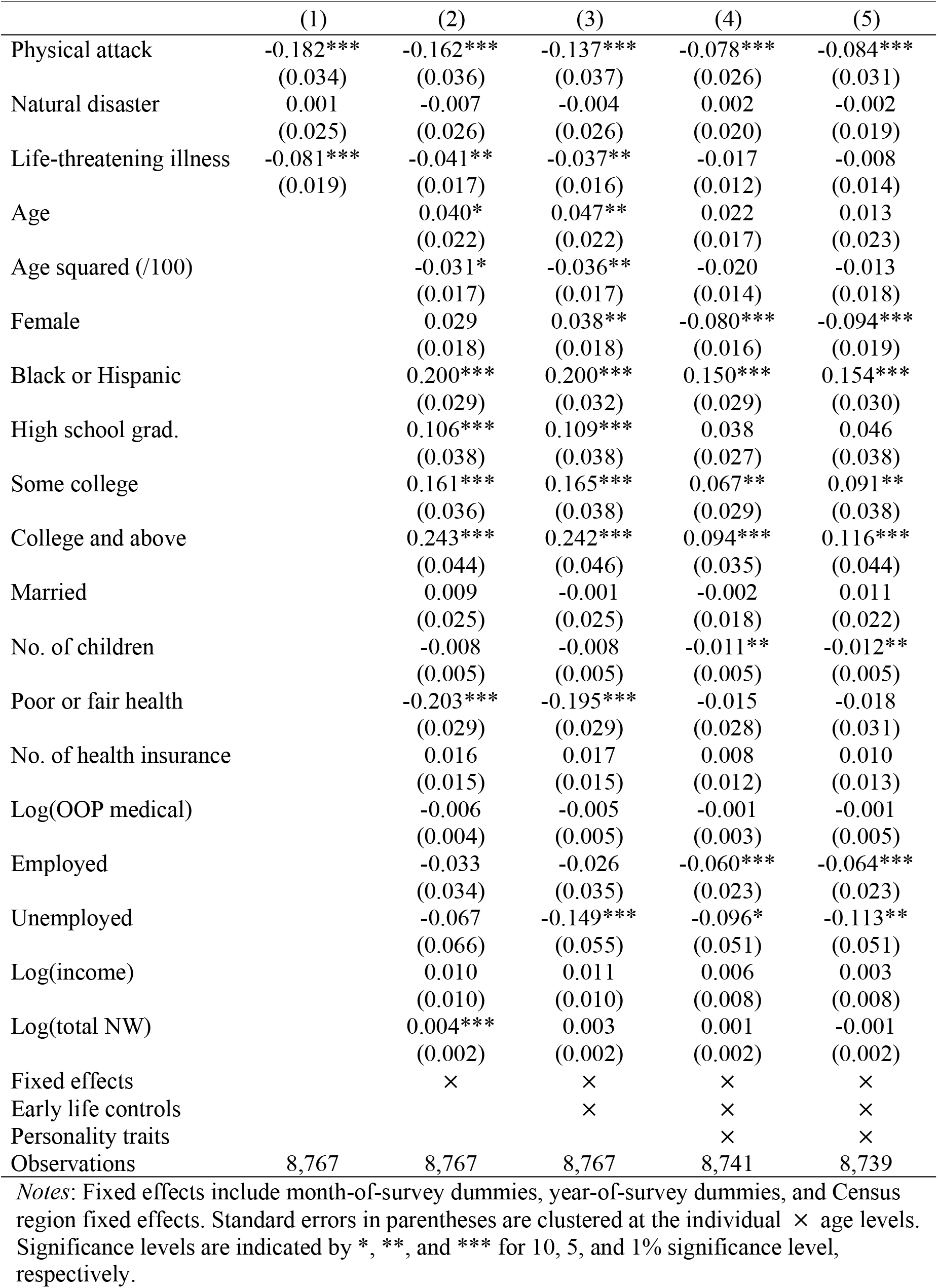
Self-control and traumatic experiences

|  | (1) | (2) | (3) | (4) | (5) |
| --- | --- | --- | --- | --- | --- |
| Physical attack | -0.182***<br>(0.034) | -0.162***<br>(0.036) | -0.137***<br>(0.037) | -0.078***<br>(0.026) | -0.084***<br>(0.031) |
| Natural disaster | 0.001<br>(0.025) | -0.007<br>(0.026) | -0.004<br>(0.026) | 0.002<br>(0.020) | -0.002<br>(0.019) |
| Life-threatening illness | -0.081***<br>(0.019) | -0.041**<br>(0.017) | -0.037**<br>(0.016) | -0.017<br>(0.012) | -0.008<br>(0.014) |
| Age |  | 0.040*<br>(0.022) | 0.047**<br>(0.022) | 0.022<br>(0.017) | 0.013<br>(0.023) |
| Age squared (/100) |  | -0.031*<br>(0.017) | -0.036**<br>(0.017) | -0.020<br>(0.014) | -0.013<br>(0.018) |
| Female |  | 0.029<br>(0.018) | 0.038**<br>(0.018) | -0.080***<br>(0.016) | -0.094***<br>(0.019) |
| Black or Hispanic |  | 0.200***<br>(0.029) | 0.200***<br>(0.032) | 0.150***<br>(0.029) | 0.154***<br>(0.030) |
| High school grad. |  | 0.106***<br>(0.038) | 0.109***<br>(0.038) | 0.038<br>(0.027) | 0.046<br>(0.038) |
| Some college |  | 0.161***<br>(0.036) | 0.165***<br>(0.038) | 0.067**<br>(0.029) | 0.091**<br>(0.038) |
| College and above |  | 0.243***<br>(0.044) | 0.242***<br>(0.046) | 0.094***<br>(0.035) | 0.116***<br>(0.044) |
| Married |  | 0.009<br>(0.025) | -0.001<br>(0.025) | -0.002<br>(0.018) | 0.011<br>(0.022) |
| No. of children |  | -0.008<br>(0.005) | -0.008<br>(0.005) | -0.011**<br>(0.005) | -0.012**<br>(0.005) |
| Poor or fair health |  | -0.203***<br>(0.029) | -0.195***<br>(0.029) | -0.015<br>(0.028) | -0.018<br>(0.031) |
| No. of health insurance |  | 0.016<br>(0.015) | 0.017<br>(0.015) | 0.008<br>(0.012) | 0.010<br>(0.013) |
| Log(OOP medical) |  | -0.006<br>(0.004) | -0.005<br>(0.005) | -0.001<br>(0.003) | -0.001<br>(0.005) |
| Employed |  | -0.033<br>(0.034) | -0.026<br>(0.035) | -0.060***<br>(0.023) | -0.064***<br>(0.023) |
| Unemployed |  | -0.067<br>(0.066) | -0.149***<br>(0.055) | -0.096*<br>(0.051) | -0.113**<br>(0.051) |
| Log(income) |  | 0.010<br>(0.010) | 0.011<br>(0.010) | 0.006<br>(0.008) | 0.003<br>(0.008) |
| Log(total NW) |  | 0.004***<br>(0.002) | 0.003<br>(0.002) | 0.001<br>(0.002) | -0.001<br>(0.002) |
| Fixed effects |  | × | × | × | × |
| Early life controls |  |  | × | × | × |
| Personality traits |  |  |  | × | × |
| Observations | 8,767 | 8,767 | 8,767 | 8,741 | 8,739 |

*Notes:* Fixed effects include month-of-survey dummies, year-of-survey dummies, and Census region fixed effects. Standard errors in parentheses are clustered at the individual × age levels. Significance levels are indicated by \*, \*\*, and \*\*\* for 10, 5, and 1% significance level, respectively.

The null model in column 1 shows a negative correlation of self-control with experience of a physical attack (*β*= − 0.182; *p* < 0.01) and a life-threatening illness (*β*= − 0.081; *p* < 0.01). After we adjust for the baseline covariates and fixed effects (column 2), the coefficient estimates on the two trauma indicators are lower but remain negatively significant at the 5% level. A rough comparison to the sample mean of self-control shows, on average, a 3.7% decline (= − 0.162 / 4.36) and a 0.94% decline (= − 0.041 / 4.36) for the experience of an attack and illness, respectively. Further conditioning on early life conditions (column 3) leads to a coefficient estimate of − 0.137, or a 3.1% decline in self-control in response to an attack (= − 0.137 / 4.36), and an estimate of − 0.037, or a 0.8% decline in self-control due to illness (= − 0.037 / 4.36), which remains statistically significant at the 5% level. The estimate of a 3.1% decline is roughly comparable to the effect of being unemployed and is half the effect of having a college degree or being an ethnic minority. The fact that a memory of almost 30 years ago explains as much as the influence of education and race bolsters our hypothesis that the influence of traumatic experience is long-lasting.

The regression in column 4 additionally controls for measures reflecting the Big Five personality traits (neuroticism, extraversion, openness to experience, agreeableness, and conscientiousness). This specification allows us to test whether the results from the previous specification are driven by a shift in personality or a shift in self-control skills beyond the respondents’ personality. The results show that the estimate on physical attacks remains negatively significant (*β*= − 0.078; *p* < 0.01), while the estimate on life-threatening illnesses is no longer different from zero at the 5% level. This result suggests that the influence of physical attacks was not limited to personality shifts but led to a meaningful reduction in one’s ability to resist temptation.

The regression in column 5 is weighted by the propensity score weight from matching the sample by exposure to any traumatic event. Matching on “observables” has the advantage of balancing out differences in the observed characteristics between the treated (with trauma) and untreated (without trauma) groups, thereby reducing the bias resulting from non-random exposure to a traumatic event. We estimate a logistic regression for having been exposed to any traumatic event (one for exposure to any event and zero otherwise) and use the kernel matching method with a bandwidth of 0.05. Similar to the unmatched results, re-weighting regression using the propensity score weight leads to a negative estimate on physical attack (*β*= − 0.084; *p* < 0.01).

Next, we examine whether the association between a traumatic event and self-control differs according to the exposure timing (see Table 3). For each traumatic event, we create a dummy variable for an event that occurred at age *x* or less (early life exposure) and another for an event that occurred after age *x* (late-life exposure), where *x* is the critical age of brain maturity. The age at which the brain stops developing and reaches its peak is not clear, with recent evidence showing that the human brain continues to mature into a person’s 20s or 30s (see Somerville, 2016 for a review). The fMRI study by Dosenbach et al. (2010) has shown, for instance, that the human brain reaches its adult volume by the age of 22, while some regions like the frontal lobe (the area of the brain concerned with self-control) continue to grow past this age, until the age of 24 or 32. Currently, there is no agreed-upon threshold for full brain maturity. Therefore, we consider the three ages posited in Dosenbach et al. (2010) as the likely threshold age of brain maturity when constructing the early-and late-life trauma exposure indicators.

**Table 3.** Self-control and traumatic experiences, by timing of events

| Cutoff age: | 22 | 24 | 32 |
| --- | --- | --- | --- |
|  | (1) | (2) | (3) |
| $\alpha_{1,1}$: Physical attack ≤ cutoff age | -0.164***<br>(0.061) | -0.149**<br>(0.060) | -0.193***<br>(0.035) |
| $\alpha_{1,2}$: Physical attack $>$ cutoff age | -0.162***<br>(0.047) | -0.169***<br>(0.048) | -0.129**<br>(0.054) |
| Natural disaster ≤ cutoff age | -0.008<br>(0.058) | -0.027<br>(0.057) | -0.032<br>(0.046) |
| Natural disaster $>$ cutoff age | -0.007<br>(0.024) | -0.001<br>(0.025) | 0.006<br>(0.023) |
| Life-threatening illness ≤ cutoff age | -0.034<br>(0.045) | -0.026<br>(0.039) | -0.025<br>(0.033) |
| Life-threatening illness $>$ cutoff age | -0.042**<br>(0.019) | -0.043**<br>(0.018) | -0.045**<br>(0.018) |
| Observations | 8,767 | 8,767 | 8,767 |
| Linear restriction ($p$-value): | | | |
| $H_0: \alpha_{1,1} - \alpha_{1,2} = 0$ | 0.98 | 0.80 | 0.25 |

*Notes:* All regressions control for baseline covariates and fixed effects. Standard errors in parentheses are clustered at the individual × age levels. Significance levels are indicated by \*, \*\*, and \*\*\* for 10, 5, and 1% significance level, respectively.

The regressions in Table 3 are conditioned on the baseline covariates and fixed effects as specified in eq. (1). Across all columns, exposure to a traumatic event is negatively related to self-control regardless of when the event occurred (*p* < 0.05 for all estimates of *α*). The F-tests for the pair of estimates concerning experience at age ≤ *x* vs. experience at age > *x* fail to reject the null hypothesis at the 5% level (linear restrictions at the bottom of Table 3), indicating that the correlation between physical attacks and self-control does not differ according to the exposure timing. Overall, there is no evidence that an attack experienced at the critical age for brain development is more meaningful for self-control skills at older ages.

Finally, we evaluate the predictive validity of our self-control measure by examining its relationship with important life outcomes known to be correlated with self-control (see Table 4). Following the evidence that self-control predicts health, health behavior, and financial well-being, we estimate the association between our measure of self-control and body mass index (BMI), obesity (BMI > 30), number of chronic health conditions, depression (Center for Epidemiologic Studies Depression scale), exercise (one for moderate physical activity zero for no physical activity), log of financial net worth, log of total net worth, and life satisfaction (using the seven-point scale of Diener et al. [1985]). Similar to the previous regressions, these models all include the baseline covariates and fixed effects. O.verall, the results confirm the prior finding that self-control is positively related to health outcomes (lower BMI, less obesity, less chronic disease, and fewer depressive symptoms), health behavior (increased participation in physical activity), financial well-being (financial and total net worth), and life satisfaction. All the coefficient estimates on self-control are statistically significant at the 5% level and are large enough to be economically meaningful. Though suggestive, these results show that our self-control measure is a valid indicator of the ability to resist temptation over many domains of behavior.

**Table 4.**
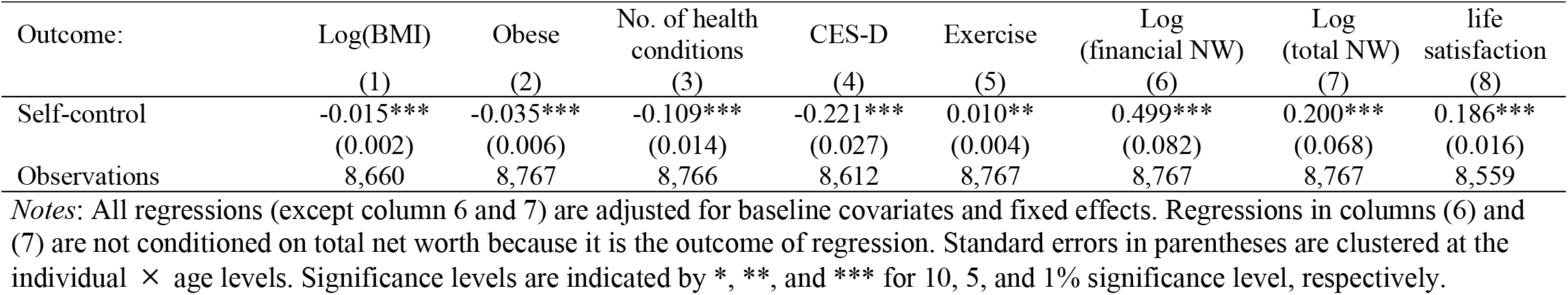
Health, well-being, and life satisfaction

| Outcome: | Log(BMI) | Obese | No. of health conditions | CES-D | Exercise | Log (financial NW) | Log (total NW) | life satisfaction |
| --- | --- | --- | --- | --- | --- | --- | --- | --- |
|  | (1) | (2) | (3) | (4) | (5) | (6) | (7) | (8) |
| Self-control | -0.015***<br>(0.002) | -0.035***<br>(0.006) | -0.109***<br>(0.014) | -0.221***<br>(0.027) | 0.010**<br>(0.004) | 0.499***<br>(0.082) | 0.200***<br>(0.068) | 0.186***<br>(0.016) |
| Observations | 8,660 | 8,767 | 8,766 | 8,612 | 8,767 | 8,767 | 8,767 | 8,559 |

*Notes:* All regressions (except column 6 and 7) are adjusted for baseline covariates and fixed effects. Regressions in columns (6) and (7) are not conditioned on total net worth because it is the outcome of regression. Standard errors in parentheses are clustered at the individual × age levels. Significance levels are indicated by \*, \*\*, and \*\*\* for 10, 5, and 1% significance level, respectively.

## 5. Discussion

The willpower to override impulses and follow through on a plan is important for individual intertemporal choice. While the behavioral implications of self-control have been studied extensively, its determinants are yet to be fully understood. This study argues convincingly that self-control can be influenced by lifetime exposure to traumatic events. Our empirical analyses have considered various potential traumatic events and have examined their associations with self-control at older ages.

Our main results reveal an average 3.1% reduction in self-control in response to a serious physical attack or assault experience. The association between exposure to attack and self-control does not differ according to whether the attack was experienced before or after age 24. The mean of the years elapsed since the attack or assault is about 30, conditional on exposure, meaning that self-control skills never fully recover to their pre-exposure level. For other types of traumatic events (natural disasters and life-threatening illnesses), we have found no consistent evidence that self-control shifts in response to exposure. Our inference regarding the effects of physical attacks and assaults is robust to controlling for early life conditions and using the propensity score re-weighting to balance out underlying heterogeneity by trauma exposure.

The weak correlations of self-control with natural disasters and life-threatening illnesses might best be explained with reference to the strength of the emotional backlash each event triggers. It is well-known that events that are more shocking and salient are more durable and accessible in memory. Among the three events considered in our study, physical attacks and assaults may be more emotionally intense given their criminal nature. Though suggestive, our preliminary findings have revealed a strongly positive correlation between physical attacks or assaults and the number of depressive symptoms. This raises the possibility that attacks and their related emotional changes manifest as post-traumatic stress disorder and disrupt the brain’s modulatory core.

Our study has several limitations. First, the higher mortality in the trauma group raises the concern that our sample may under-represent severely traumatized individuals and over-represent those exposed to relatively low risk (Pak, 2021). Under the plausible assumption that those excluded from the sample are the most impulsive, addressing survivorship bias would lead to a stronger negative correlation between trauma exposure and self-control. Second, it remains unclear through which mechanism traumatic experience produces a lasting influence on self-control. Our estimates could be the result of mental health problems such as post-traumatic stress disorder or its influence through employment outcomes or portfolio reallocation. Future research needs to examine cohort data that could track a lifetime transition into old age and explore the various channels underlying the consequences of trauma. Finally, individuals might differ in terms of their propensity to experience traumatic events. Although our childhood covariates and matching estimation mitigate these effects, we cannot completely rule out the possibility that some aspects of mid-life are still unobserved and confound our estimation process. Finally, we were unable to identify a causal relationship between traumatic experience and self-control. It would be fruitful to exploit exogenous changes in traumatic experiences, such as natural disasters and armed conflicts, and examine their correlation with self-control outcomes at older ages.

Despite these limitations, our findings offer a useful guide for future researchers examining the formation of self-control skills. Specifically, it is important to note that the demographics, socioeconomic characteristics, and trauma indicators used in this study jointly explain less than 40% of an individual’s self-control. The limited explanatory power of the estimated models calls for a more rigorous and systematic investigation into how individual self-control is determined. Researchers in behavioral economics are encouraged to replicate or expand this study using multiple self-control assessments, different sample frames, and various domains of self-control, in both the experimental setting and the survey context.

Among its key implications, this study highlights the need for government intervention to treat traumatic experiences. While mental health programs are available for war veterans, those outside the military are often overlooked and are not provided with targeted intervention. As discussed above, having an ability to resist temptation has broad benefits for individuals, their family, and society they belong to. Our findings imply that a well-designed mental health program that explicitly focuses on traumatized individuals and provides support early in life would produce significant social benefits. Since those at the bottom of the economic scale are more likely to experience traumatic events, the benefits of such policy measures would disproportionately accrue to the least advantaged social groups.

## Data Availability

All data files are available from the Health and Retirement Study database.

---

**Table A1.** Description of variables

| Variable | Survey questions and coding |
| --- | --- |
| Trauma variables: |  |
| Physical attack | “Were you the victim of a serious physical attack or assault in your life?” |
| Natural disaster | “Have you even been in a major fire, flood, earthquake, or other natural disaster?” |
| Life-threatening illness | “Did you ever have a life-threatening illness or accident?” |
| Baseline covariates: |  |
| Age | Age at the time of survey |
| Female | Female = 1; male = 0 |
| Black or Hispanic | African American or Hispanic = 1; White or other races = 0 |
| High school graduate | Years of schooling = 12 |
| Some college | Years of schooling = 13, 14, or 15 |
| College and above | Years of schooling ≥ 16 |
| Married | Married = 1; separated, divorced, widowed, or never married = 0 |
| No. of living children | Number of living children |
| Poor or fair health | Self-rated health: poor or fair = 1; good, very good, or excellent = 0 |
| No. of health insurance | Count of employer-provided / private health insurance plans |
| OOP medical spending | OOP medical expenditures since the last interview |
| Employed | Works full-time or part-time = 1; unemployed or retired = 0 |
| Unemployed | Unemployed = 1; works full-time or part-time or retired = 0 |
| Household income | Sum of household earnings, pensions and annuities, Social Security benefits, government transfers, capital income, and other income |
| Total net worth | Net value of checking and savings accounts, bonds and bond funds, stocks and stock mutual funds, retirement accounts, primary home, real estate, business equity, vehicles, and other savings |
| Early-life characteristics: |  |
| Place of birth | Born in South Atlantic, West South Central, and East South Central = 1; other regions = 0 |
| Mother’s education background | Mother’s years of education |
| Father’s education background | Father’s years of education |
| Quality of relationship with mother | “I had a good relationship with my mother before age 18” (Agree or strongly agree = 1; neither agree nor disagree, disagree, or strongly disagree = 0) |
| Quality of relationship with father | “I had a good relationship with my father before age 18” (Agree or strongly agree = 1; neither agree nor disagree, disagree, or strongly disagree = 0) |
| Substance abuse of parents | “Before you were 18 years old, did either of your parents drink or use drugs so often that it caused problems in the family?” (yes = 1; no = 0) |
| OOP medical spending (earlier wave) | OOP medical expenditures recorded in the 1998 wave (HRS, AHEAD, CODA, and WB cohorts); OOP medical expenditures recorded in the 2004 wave (EBB cohort) |
| Employed (earlier wave) | Employed in 1998 (HRS, AHEAD, CODA, and WB cohorts); employed in 2004 (EBB cohort) |
| Unemployed (earlier wave) | Unemployed in 1998 (HRS, AHEAD, CODA, and WB cohorts); unemployed in 2004 (EBB cohort) |
| Household income (earlier wave) | Household income in 1998 (HRS, AHEAD, CODA, and WB cohorts); household income in 2004 (EBB cohort) |
| Total net worth (earlier wave) | Total net worth in 1998 (HRS, AHEAD, CODA, and WB cohorts); total net worth in 2004 (EBB cohort) |

